# Diagnosing Sepsis Through Proteomic Insights: Findings from a Prospective ICU Cohort

**DOI:** 10.1101/2025.08.26.25334458

**Authors:** A Khaleghi Ardabili, S Rice, AS Bonavia

## Abstract

**Introduction:** Sepsis diagnosis remains clinical and heterogeneous. We hypothesized that a proteomics-informed machine-learning approach could identify a small, easy-to-use, and optimized set of clinical variables to complement or potentially outperform SOFA.

**Methods:** We conducted a prospective, single-center, observational study in an academic intensive care unit. Plasma from critically ill patients with and without sepsis was analyzed using liquid chromatography coupled with tandem mass spectrometry (LC-MS). Data were acquired with data-independent acquisition parallel accumulation– serial fragmentation (diaPASEF) and processed using DIA-NN software. Differentially expressed proteins informed model development. Random Forest models were trained in a Discovery cohort (n=55) to select clinical variables linked to the proteome, then tested in an independent Validation cohort (n=59). Recursive feature elimination (RFE) identified a minimal feature set that was predictive of sepsis. The performance was assessed using repeated cross-validation and external validation.

**Results:** Twelve plasma proteins differed between sepsis and non-sepsis patients at FDR < 0.1, corresponding to 26 proteome-enriched clinical variables. The classifier achieved mean AUC’s of 0.73 and 0.76 in Discovery and Validation cohorts, respectively. RFE performance plateaued with ≥9 variables, peaked at an accuracy of 0.78, and deteriorated below seven; the final three features before collapse were plasma BUN, chemokine ligand 3 (CCL3), and creatinine. Proteome-to-clinical regression highlighted creatinine as having the strongest correlation (R² = 0.558). **Discussion**: A concise set of routinely obtainable variables anchored by renal markers and CCL3 captured proteomic signals and discriminated sepsis across cohorts, supporting a “proteomics-informed, clinic-first” strategy for pragmatic EHR deployment.

While larger multicenter studies are warranted, these findings suggest that renal dysfunction exerts a disproportionate influence on sepsis and that increased emphasis on kidney-related markers may improve both recognition and risk assessment.

## Introduction

Sepsis is a clinical syndrome characterized by organ dysfunction, resulting from a dysregulated host immune response to infection. Despite decades of investigation, the clinical and immunological heterogeneity of sepsis has prevented the identification of a single biomarker that is pathognomonic for this condition. Consequently, the diagnosis remains clinical and is based on consensus definitions alone (1).

Organ dysfunction in sepsis is operationalized using the Sequential (or Sepsis-Related) Organ Failure Assessment (SOFA) score, which predates the current sepsis definitions and was not originally developed as a diagnostic tool for the syndrome (2). The SOFA score is a composite measure of dysfunction across six equally weighted organ systems: respiratory, coagulation, hepatic, cardiovascular, central nervous, and renal. It is designed to be easily used in the clinical setting, facilitating the rapid identification of patients with sepsis for timely medical management. Under the Sepsis-3 Criteria, a diagnosis of sepsis requires both suspected infection and an increase in SOFA score of two points or more from baseline, independent of the organ system(s) involved.

We hypothesized that machine-learning models could identify clinical predictors of sepsis in a data-driven yet easy-to-use manner. By integrating high-dimensional proteomic data with established clinical measures, we aimed to reveal complex nonlinear associations that might elude conventional statistical approaches. Clinical data utilized in the discovery process included all variables that form the foundation of the SOFA score, together with other laboratory values, physiological parameters, and biomarkers that are also commonly measured in critically ill patients. A similar strategy was successfully employed by Pimienta et al., who used quantitative mass spectrometry-based proteomics to analyze plasma from murine sepsis models, identifying a functionally relevant network of extracellular proteins, termed the plasma proteasome signature of sepsis (3).

Given the current impracticality of comprehensive proteomic sequencing in clinical settings, our goal was to leverage plasma proteomic profiles to distinguish sepsis from non-sepsis critical illnesses as a foundation for identifying clinically measurable variables (Discovery Cohort). We aimed to validate the predictive value of these empirically derived clinical variables in an independent patient cohort (Validation Cohort). This approach capitalizes on the abundance and accessibility of clinical data to develop rapid, practical tools for the early identification of rapidly progressive diseases, such as sepsis.

## Materials and Methods

### Study Design and Participants

This prospective observational study was conducted at an academic medical center between October 2021 and July 2024. The study adhered to the STROBE guidelines (4) and was approved by the institutional review board. Eligible participants were adults admitted to the intensive care unit (ICU) who met the criteria for critical illness and suspected sepsis. They were identified using an electronic health record algorithm based on Modified Early Warning Scores (MEWS). Two independent clinicians reviewed all flagged cases to confirm their eligibility. Informed consent was obtained from the patients or their legally authorized representatives.

### Eligibility

Adults (≥18 years) enrolled within 48 h of critical illness development were included. Sepsis was diagnosed based on the Sepsis-3 criteria (5, 6), defined as an increase in SOFA score ≥2 points associated with suspected or confirmed infection. A diagnosis of critical illness required respiratory support or vasopressor therapy in an ICU setting. Patients were excluded for conditions known to impact immune function, including active blood cancers, immunosuppressive treatments, chronic corticosteroid use (unless sepsis-related), recent chemotherapy or radiation therapy, HIV infection with low CD4 counts, solid organ transplantation, or autoimmune diseases, such as lupus or rheumatoid arthritis.

### Data Collection

Clinical data, including patient demographics, measures of disease severity, laboratory results, and infection characteristics, were obtained from an institutional critical illness repository (**Supplementary Table S1**). These features encompassed a combination of previously described and commonly cited characteristics that are used to describe sepsis and critical illness. Comorbidities were assessed using the Charlson Comorbidity Index (7), and illness severity was measured using the APACHE II score (8–10). Shock was defined as elevated lactate level (>2 mmol/L) despite adequate fluid resuscitation and vasopressor use.

### Blood Collection and Processing

Blood samples for proteomic analyses were collected in EDTA tubes, centrifuged, and plasma was frozen at-80°C. Plasma was later thawed and processed as a single batch; thawing was supplemented with protease inhibitors, and proteins were extracted and digested using automated magnetic-bead-based methods on a KingFisher Flex platform.

### Proteomic Analysis

Proteins were analyzed by liquid chromatography-mass spectrometry (LC-MS) using data-independent acquisition parallel accumulation–serial fragmentation (dia-PASEF). Data were processed using DIA-NN (Aptila Biotech GmbH, Berlin, Germany) to identify and quantify proteins.

Differentially expressed proteins between sepsis and non-sepsis groups were identified using Ingenuity Pathway Analysis (QIAGEN Silicon Valley, Redwood City, CA, USA).

### Statistical Analysis and Machine Learning

We used Random Forest models to identify clinical variables that could be accurately predicted from proteomic profiles (Discovery Cohort, n=55). Continuous clinical variables (such as age, lactate levels, and SOFA scores) and binary variables (such as sex) were tested. Variables that were reliably predicted by proteomics were then tested for their ability to predict the diagnosis of sepsis. The best-performing clinical variables were further validated in an independent Validation Cohort (n=59). **Fig 1** illustrates the experimental workflow from sample collection to bioinformatic analysis.

**Fig 1.**
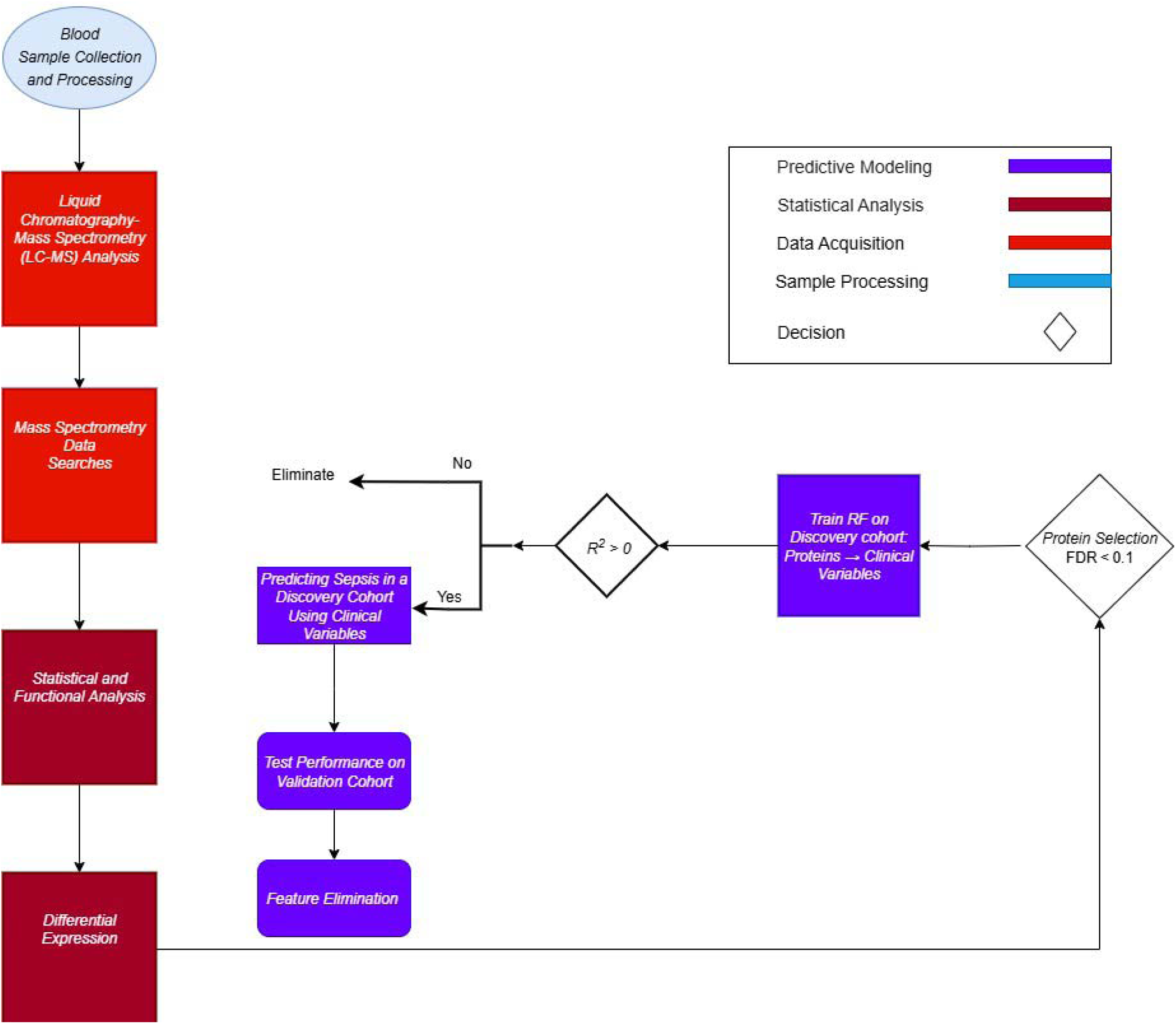
**Flow diagram summarizing the proteomics-informed predictive modeling approach**. Blood samples from critically ill patients underwent sample processing and proteomic data acquisition using Liquid Chromatography-Mass Spectrometry (LC-MS). Differentially expressed proteins between sepsis and non-sepsis patients were identified. These proteins (FDR < 0.1) were then used in Random Forest predictive modeling to predict clinical variables in a Discovery cohort (n = 55). Clinical variables successfully predicted by proteomic profiles were retained and subsequently used to predict sepsis status in an independent Validation cohort (n = 59). Recursive Feature Elimination further identified a minimal set of the most predictive clinical variables.

## Results

### Patient Characteristics

The Discovery cohort comprised 38 sepsis and 17 critically ill but non-sepsis (CINS) patients; mean age 65.9 ± 17.2 years, mean APACHE II score 22 ± 8.1, and 40% female. The Validation cohort included 31 sepsis and 28 CINS patients with comparable acute illness severity (APACHE II 21.1 ± 6.7; p = 0.52) and sex distribution (37 % female, **Table 1**).

**Table 1.** Comparison between Discovery and Validation Cohorts.

|  | Discovery Cohort | Validation Cohort |
| --- | --- | --- |
| Sepsis (n, %) | 40 (71.4%) | 31 (52.5%) |
| Age, years | 70.5 [60.0, 76.0] | 70.0 [61.0, 77.5] |
| APACHE II score | 22.9 ± 8.0 | 21.1 ± 6.7 |
| Lactate, plasma (mmol/dl) | 3.1 [2.0, 5.0] | 2.10 [0.8, 3.9] |
| Shock (n, %) | 29 (51.8%) | 50 (84.7%) |
| Absolute lymphocyte count (K/μl) | 0.75 [0.46, 1.06] | 0.99 [0.78, 1.15] |
| SOFA (day 1 of illness) | 8.0 [6.8, 11.0] | 6.0 [4.5, 10.0] |
| SOFA (maximum score) | 9.0 [6.8, 11.0] | 7.0 [5.0, 10.5] |
| CCL3, plasma (pg/ml) | 99.4 [54.6, 133.0] | 55.0 [33.6, 78.1] |
| CXCL10, plasma (pg/ml) | 379.0 [164.0, 767.0] | 213.0 [118.5, 428.0] |
| TNFR1, plasma (ng/ml) | 5.3 [3.6, 8.6] | 4.0 [2.2, 7.9] |
| Temperature, deg C | 36.3 [36.0, 38.2] | 36.2 [35.9, 38.1] |
| Diastolic blood pressure, mm Hg | 44.0 [38.0, 49.0] | 46.00 [41.0, 51.0] |
| Ventilator (n, %) | 23 (41.1%) | 31 (52.5%) |
| Glasgow Coma Score | 11.5 [6.8, 15.0] | 11.0 [7.0, 15.0] |
| LPS-stimulated IL-6 production (ng/ml) | 115.8 [39.0, 328.5] | 348.9 [188.6, 743.0] |
| LPS-stimulated TNF production (ng/ml) | 43.1 [14.9, 88.5] | 76.2 [35.5, 147.9] |
| BUN, plasma (mg/dl) | 29.0 [18.0, 47.8] | 25.0 [16.0, 43.5] |
| Albumin, plasma (g/dl) | 2.79 ± 0.57 | 3.03 ± 0.68 |
| Total bilirubin, plasma (mg/dl) | 1.20 [0.60, 2.08] | 0.90 [0.40, 1.60] |
| Creatinine, plasma (mg/dl) | 1.48 [1.01, 2.35] | 1.46 [0.86, 2.58] |
| Hematocrit, % | 31.4± 5.5 | 31.5 ± 6.1 |

### Proteome Depth and Quality

LC-MS identified 5132 precursors mapping to 696 protein groups. After stringent filtering, 507 proteins were consistently quantified, with 90% of samples displaying a median coefficient of variation < 20 % (**Supplementary Fig S1**).

### Differential Expression

Twelve proteins were differentially expressed between sepsis and CINS patients at FDR < 0.1 (**Table 2**). A heat map (**Fig 2A**) demonstrates a clear separation of groups, while a volcano plot (**Fig 2B**) highlights both the magnitude and significance of change.

**Fig 2.**
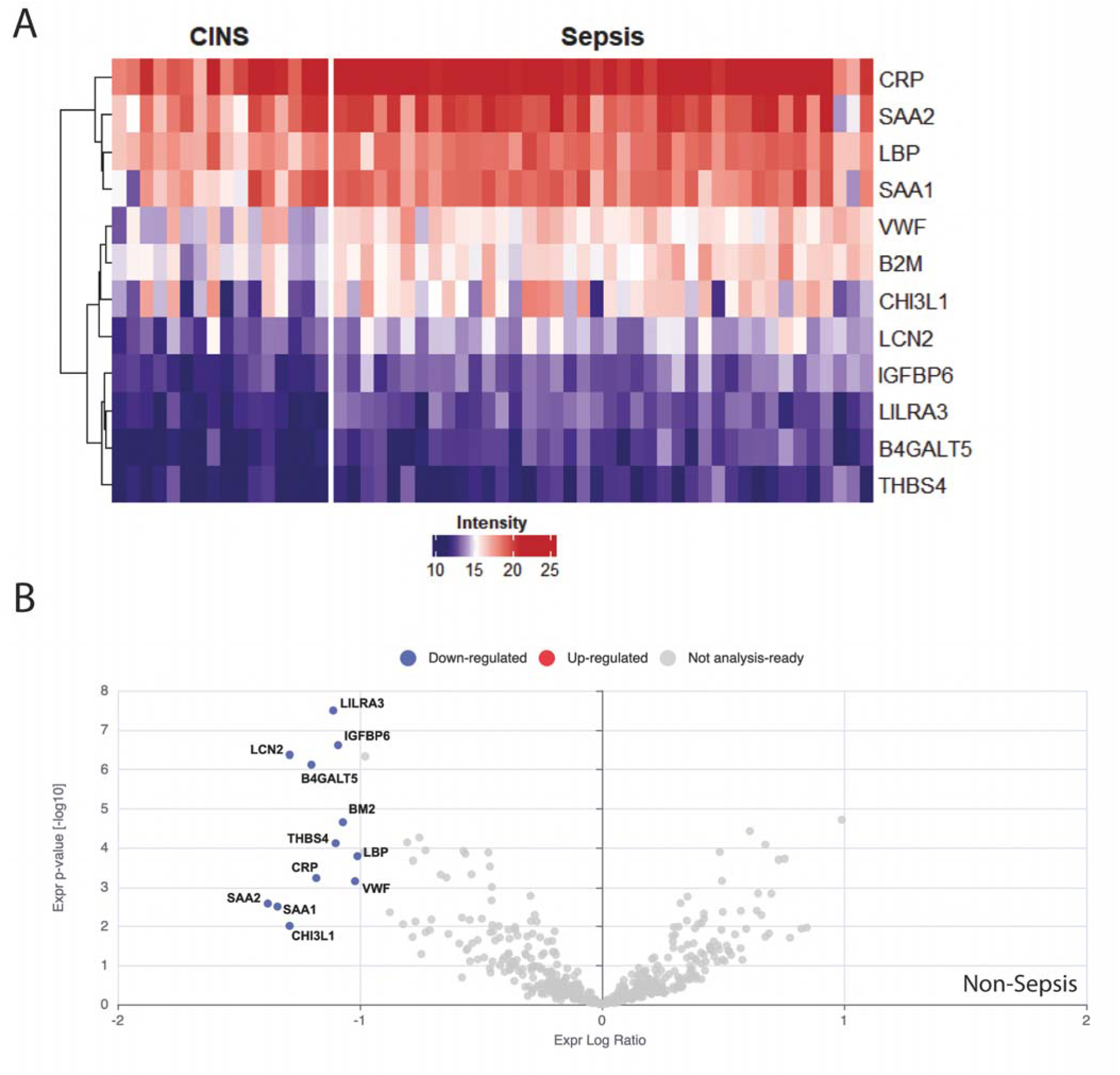
The Plasma Proteome Signature Associated with Acute Sepsis. (A) Hierarchical clustering of Discovery cohort plasma samples (columns) and 12 differentially expressed proteins (rows; FDR < 0.1). Log2-scaled intensities are *z-* standardized per protein. Sepsis samples exhibit uniformly higher expression (red) of acute-phase proteins (e.g., SAA1/2, CRP) and immune mediators (LCN2, LILRA3) relative to critical-illness non-sepsis (CINS) controls. (B) Volcano plot of differential plasma protein expression. Each dot represents a quantified protein (n = 507). The x-axis shows log2 fold-change (sepsis vs CINS); the y-axis shows –log10 adjusted p-value. Proteins meeting FDR < 0.10 and |log2-fold change| > 1 are colored red (up-regulated) or blue (down-regulated). B2M = beta-2-microglobulin, B4GALT = beta-1,4-galactosyltransferase 5, CHI3L1 = chitinase 3 like 1, CRP = C-reactive protein, IGFBP6 = insulin-like growth factor binding protein 6, LBP = liposaccharide-binding protein, LCN2 = lipocalin 2, LILRA3 = leukocyte immunoglobulin-like receptor 3, SAA1 = serum amyloid A1, SAA2 = serum amyloid A2, THBS4 = thrombospondin 4, VWF = Von Willebrand Factor.

**Table 2.** Protein expression levels in critically ill, non-septic patients relative to those in septic patients.

| Protein ID | Gene Name | Log2-fold change | *P-value* | *Bonferroni-adjusted*<br>*P-value* |
| --- | --- | --- | --- | --- |
| P0DJI9 | SAA2 | -1.38 | 0.0026 | 0.035 |
| P0DJI8 | SAA1 | -1.34 | 0.0031 | 0.041 |
| P36222 | CHI3L1 | -1.29 | 0.0097 | 0.082 |
| P80188 | LCN2 | -1.29 | $4.3 \times 10^{-7}$ | $6.0 \times 10^{-5}$ |
| O43286 | B4GALT5 | -1.20 | $7.6 \times 10^{-7}$ | $7.7 \times 10^{-5}$ |
| P02741 | CRP | -1.18 | $5.8 \times 10^{-4}$ | 0.010 |
| Q8N6C8 | LILRA3 | -1.11 | $3.2 \times 10^{-8}$ | $1.6 \times 10^{-5}$ |
| P35443 | THBS4 | -1.10 | $7.7 \times 10^{-5}$ | 0.0032 |
| P24592 | IGFBP6 | -1.09 | $2.5 \times 10^{-7}$ | $6.0 \times 10^{-5}$ |
| P61769 | B2M | -1.07 | $2.2 \times 10^{-5}$ | 0.0016 |
| P04275 | VWF | -1.02 | $7.0 \times 10^{-4}$ | 0.012 |
| P18428 | LBP | -1.01 | $1.7 \times 10^{-4}$ | 0.0042 |

### Proteome-to-clinical Prediction

Of the clinical features obtained from the Critical Illness Biorepository, serum procalcitonin, C-reactive protein, and infection type were excluded owing to missing values (73%, 63%, and 29% missingness, respectively). Variables with a low percentage of missingness were imputed using multivariate imputation by chained equations (MICE).

Random forest regression of the 12 differentially expressed proteins predicted continuous clinical variables with R² > 0, notably creatinine (R² = 0.558), PD-L1 (0.387), and IL-8 (0.363) levels (Supplementary Table S2).

### Sepsis Classification Models

Using the protein-enriched clinical variable set (26 features), the model achieved a mean AUC of 0.73 ± 0.01 in the Discovery cohort, based on 10 repeated cross-validations (**Fig 3A**) and a mean AUC of 0.76 ± 0.01 in the independent Validation cohort (**Fig 3B**).

**Fig 3.**
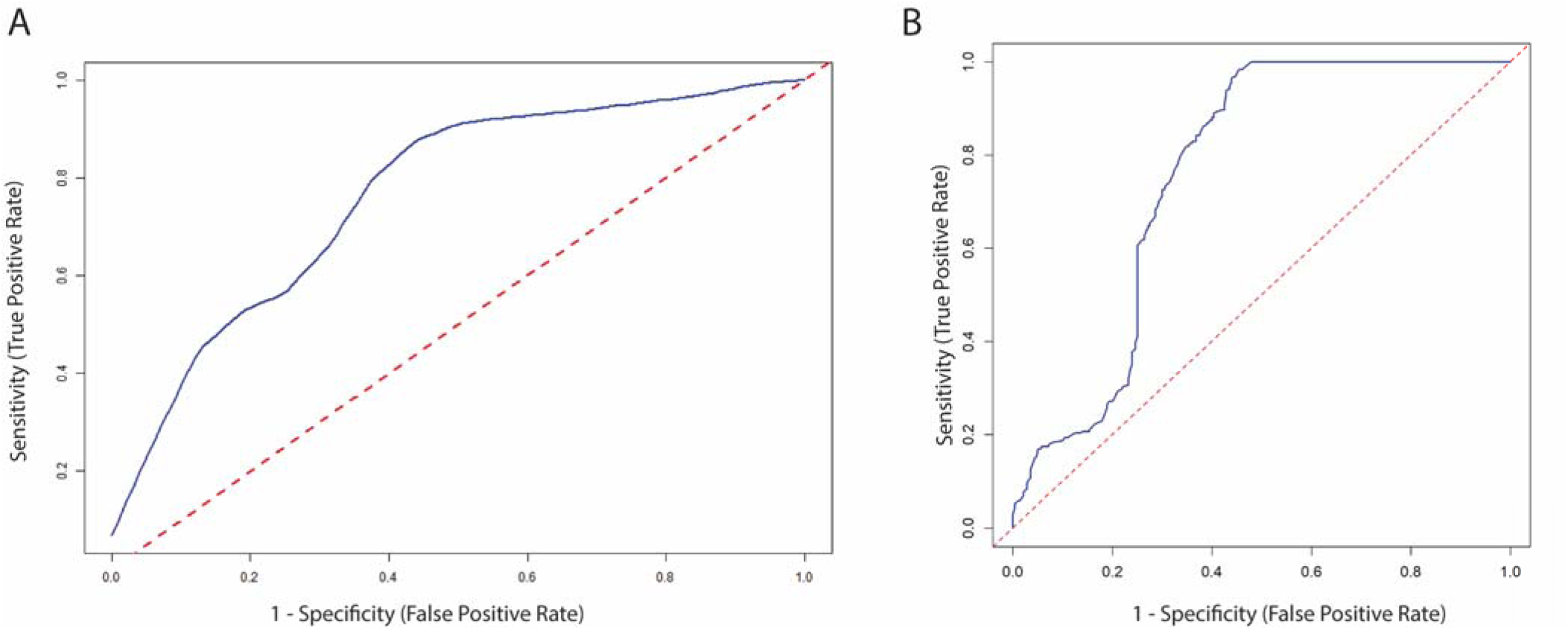
Receiver-operating Characteristic Curve for Sepsis Diagnosis. (A) Discovery cohort - The Random Forest model trained on 26 proteome-enriched clinical variables was evaluated with leave-one-out cross-validation (10 repeats). Mean area under the ROC curve (AUC) = 0.73; dashed line indicates chance (AUC = 0.50). (B) Validation cohort - The model and fixed decision threshold derived from the Discovery cohort were applied to 59 new patients. Mean AUC = 0.76 over 10 repeated runs, demonstrating robust generalizability.

### Feature Minimization

Recursive feature elimination (RFE) iteratively trimmed the initial 26-variable model while tracking the cross-validated accuracy (**Fig 4**, **Table 3**). Model performance plateaued at ∼0.75 when ≥ 9 variables were retained, peaked at 0.78 after the eighth iteration, and declined once fewer than seven variables remained. The first variables, shock and mechanical ventilation *-* had the lowest feature importance scores, and their removal did not impair the accuracy. In contrast, the accuracy fell sharply once key inflammatory mediators or metabolic indicators were eliminated. The last three features preserved before model collapse were blood urea nitrogen (BUN), chemokine ligand 3 (CCL3), and creatinine, highlighting them as the core and most informative predictors of sepsis in this cohort.

**Fig 4.**
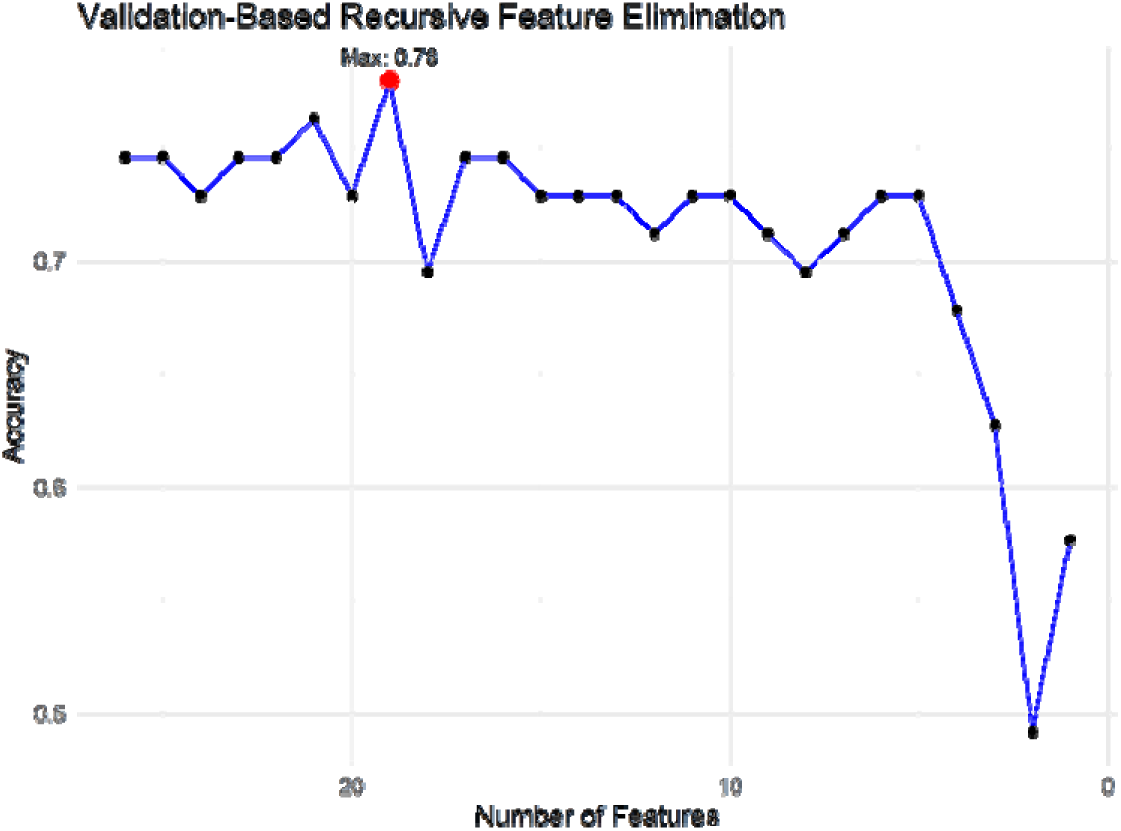
Recursive Feature Elimination Performance Curve. Cross-validated classification accuracy (y-axis) as a function of the number of retained clinical variables (x-axis). Peak accuracy is achieved with nine variables, beyond which performance plateaus, indicating an optimal minimal feature set for rapid bedside implementation.

**Table 3.**
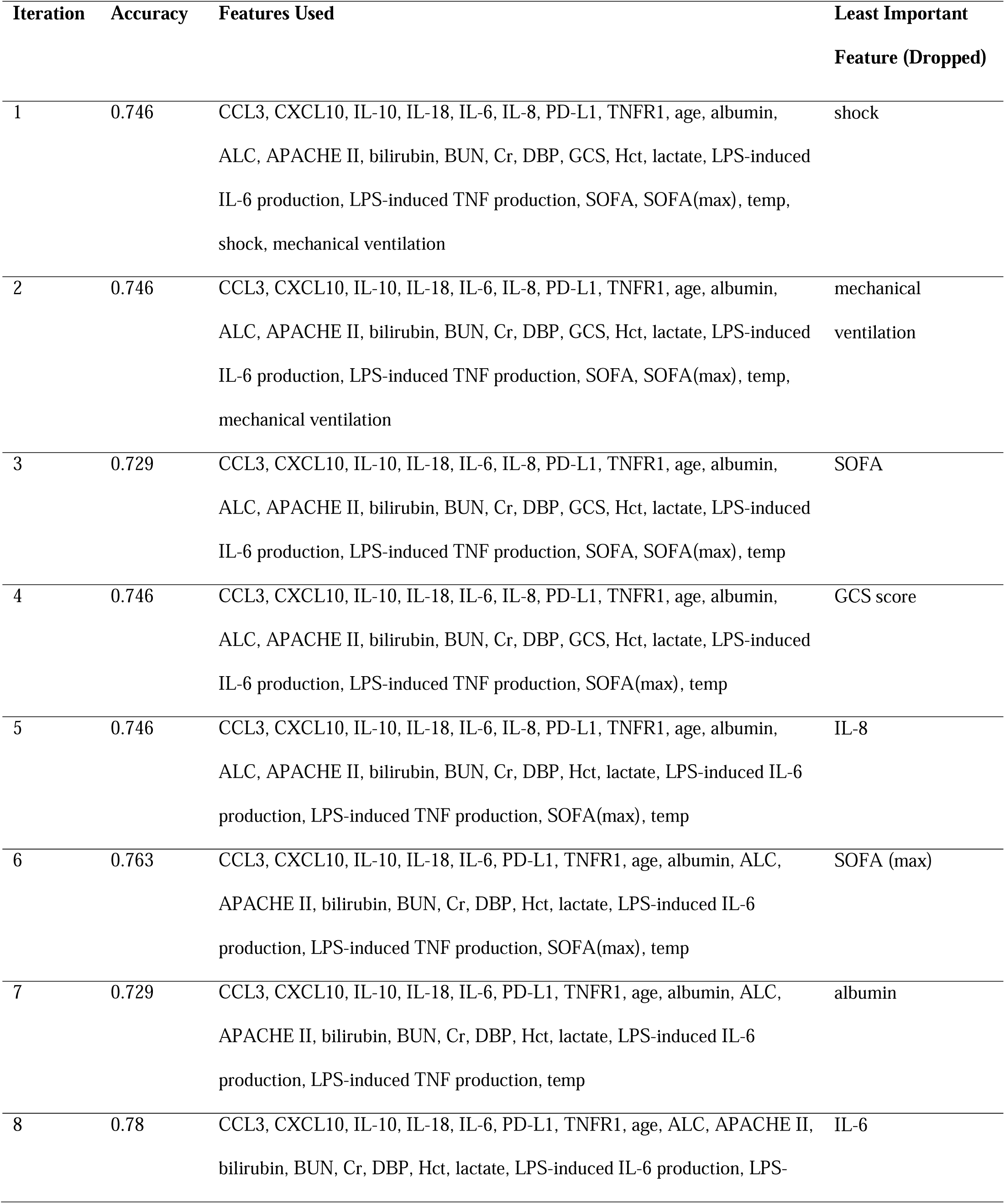
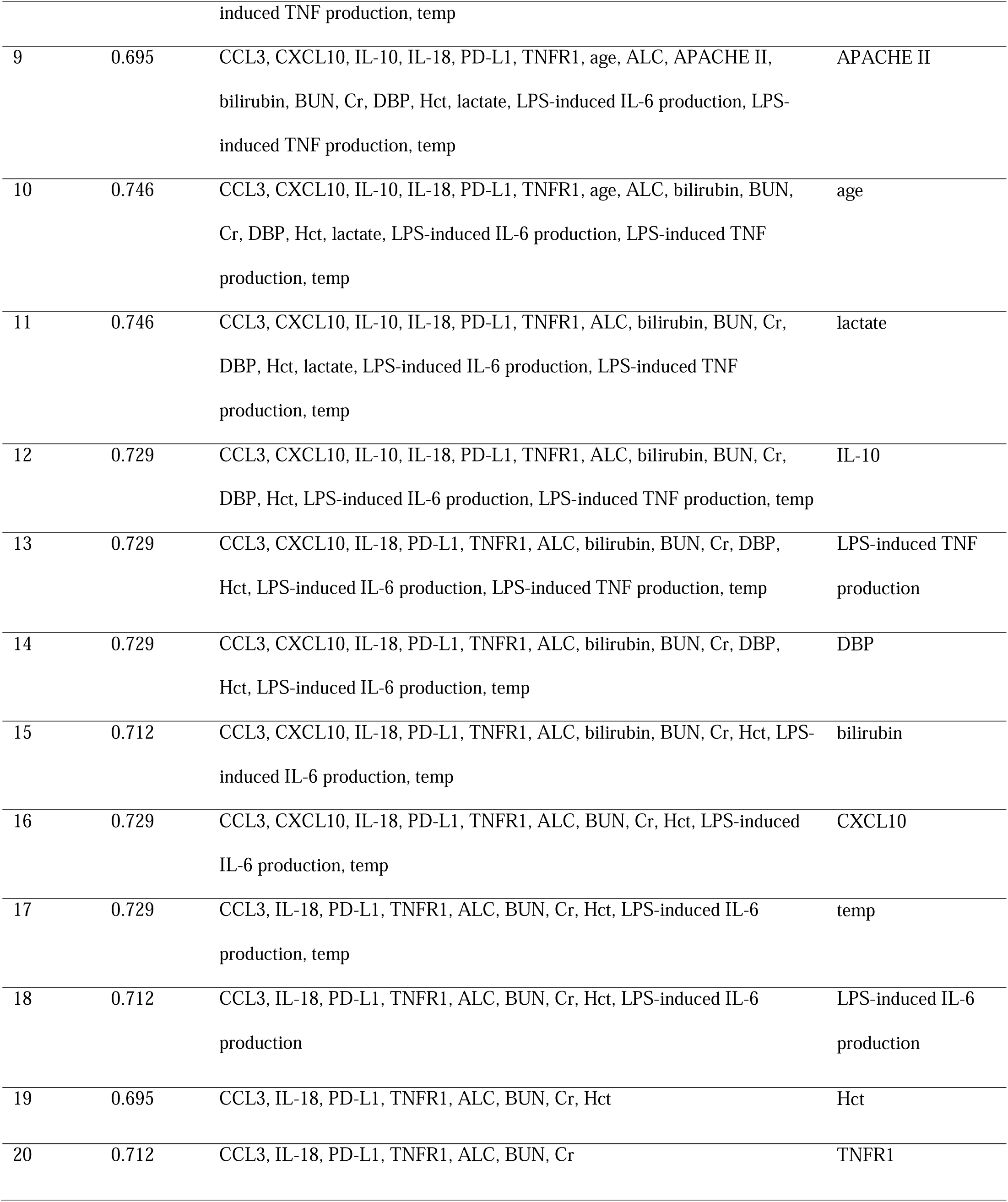
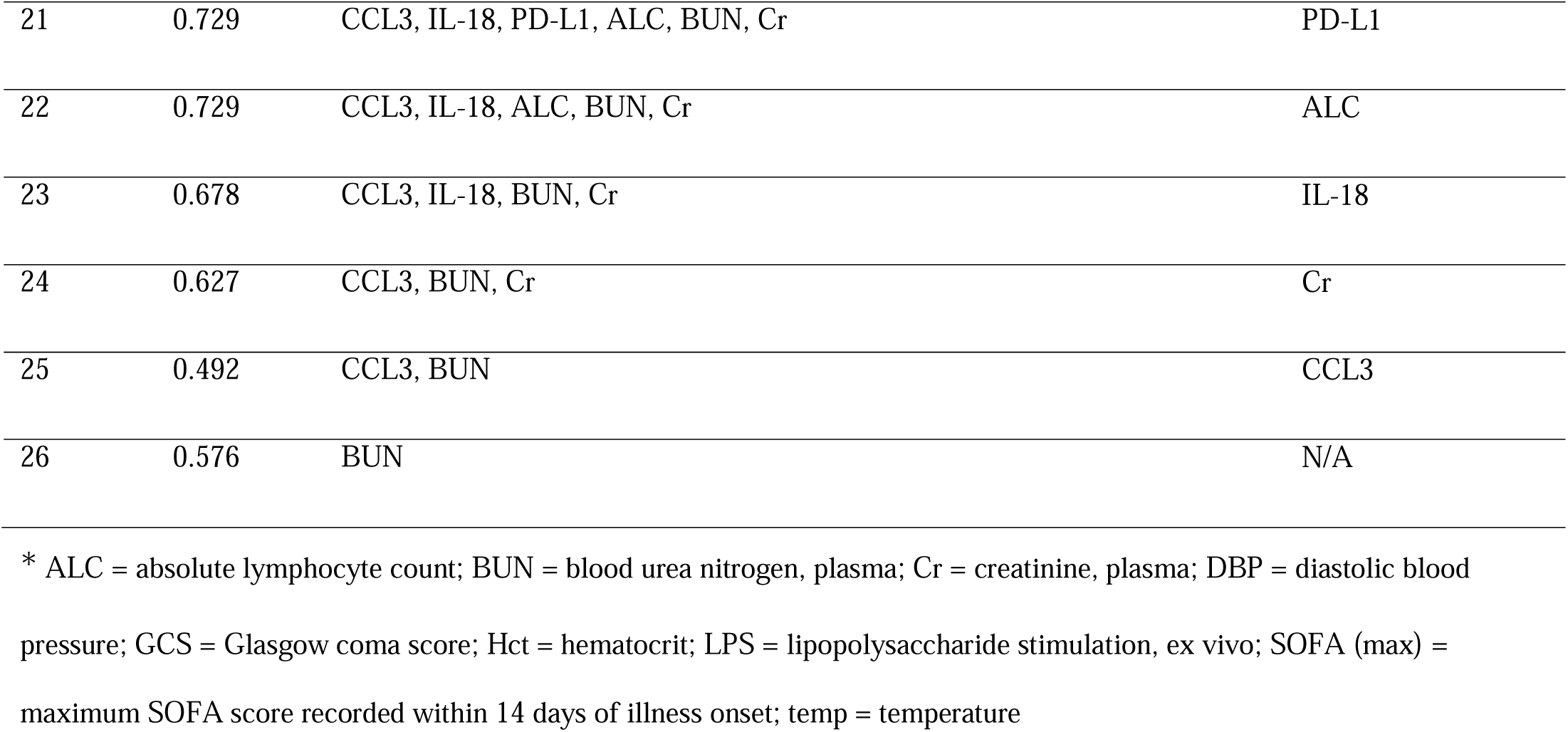
Stepwise Recursive Feature Elimination: Cross-validated accuracy, retained feature set, and least-important feature removed at each iteration.

| Iteration | Accuracy | Features Used | Least Important Feature (Dropped) |
| --- | --- | --- | --- |
| 1 | 0.746 | CCL3, CXCL10, IL-10, IL-18, IL-6, IL-8, PD-L1, TNFR1, age, albumin, ALC, APACHE II, bilirubin, BUN, Cr, DBP, GCS, Hct, lactate, LPS-induced IL-6 production, LPS-induced TNF production, SOFA, SOFA(max), temp, shock, mechanical ventilation | shock |
| 2 | 0.746 | CCL3, CXCL10, IL-10, IL-18, IL-6, IL-8, PD-L1, TNFR1, age, albumin, ALC, APACHE II, bilirubin, BUN, Cr, DBP, GCS, Hct, lactate, LPS-induced IL-6 production, LPS-induced TNF production, SOFA, SOFA(max), temp, mechanical ventilation | mechanical ventilation |
| 3 | 0.729 | CCL3, CXCL10, IL-10, IL-18, IL-6, IL-8, PD-L1, TNFR1, age, albumin, ALC, APACHE II, bilirubin, BUN, Cr, DBP, GCS, Hct, lactate, LPS-induced IL-6 production, LPS-induced TNF production, SOFA, SOFA(max), temp | SOFA |
| 4 | 0.746 | CCL3, CXCL10, IL-10, IL-18, IL-6, IL-8, PD-L1, TNFR1, age, albumin, ALC, APACHE II, bilirubin, BUN, Cr, DBP, GCS, Hct, lactate, LPS-induced IL-6 production, LPS-induced TNF production, SOFA(max), temp | GCS score |
| 5 | 0.746 | CCL3, CXCL10, IL-10, IL-18, IL-6, IL-8, PD-L1, TNFR1, age, albumin, ALC, APACHE II, bilirubin, BUN, Cr, DBP, Hct, lactate, LPS-induced IL-6 production, LPS-induced TNF production, SOFA(max), temp | IL-8 |
| 6 | 0.763 | CCL3, CXCL10, IL-10, IL-18, IL-6, PD-L1, TNFR1, age, albumin, ALC, APACHE II, bilirubin, BUN, Cr, DBP, Hct, lactate, LPS-induced IL-6 production, LPS-induced TNF production, SOFA(max), temp | SOFA (max) |
| 7 | 0.729 | CCL3, CXCL10, IL-10, IL-18, IL-6, PD-L1, TNFR1, age, albumin, ALC, APACHE II, bilirubin, BUN, Cr, DBP, Hct, lactate, LPS-induced IL-6 production, LPS-induced TNF production, temp | albumin |
| 8 | 0.78 | CCL3, CXCL10, IL-10, IL-18, IL-6, PD-L1, TNFR1, age, ALC, APACHE II, bilirubin, BUN, Cr, DBP, Hct, lactate, LPS-induced IL-6 production, LPS- induced TNF production, temp | IL-6 |
| 9 | 0.695 | CCL3, CXCL10, IL-10, IL-18, PD-L1, TNFR1, age, ALC, APACHE II, bilirubin, BUN, Cr, DBP, Hct, lactate, LPS-induced IL-6 production, LPS-induced TNF production, temp | APACHE II |
| 10 | 0.746 | CCL3, CXCL10, IL-10, IL-18, PD-L1, TNFR1, age, ALC, bilirubin, BUN, Cr, DBP, Hct, lactate, LPS-induced IL-6 production, LPS-induced TNF production, temp | age |
| 11 | 0.746 | CCL3, CXCL10, IL-10, IL-18, PD-L1, TNFR1, ALC, bilirubin, BUN, Cr, DBP, Hct, lactate, LPS-induced IL-6 production, LPS-induced TNF production, temp | lactate |
| 12 | 0.729 | CCL3, CXCL10, IL-10, IL-18, PD-L1, TNFR1, ALC, bilirubin, BUN, Cr, DBP, Hct, LPS-induced IL-6 production, LPS-induced TNF production, temp | IL-10 |
| 13 | 0.729 | CCL3, CXCL10, IL-18, PD-L1, TNFR1, ALC, bilirubin, BUN, Cr, DBP, Hct, LPS-induced IL-6 production, LPS-induced TNF production, temp | LPS-induced TNF production |
| 14 | 0.729 | CCL3, CXCL10, IL-18, PD-L1, TNFR1, ALC, bilirubin, BUN, Cr, DBP, Hct, LPS-induced IL-6 production, temp | DBP |
| 15 | 0.712 | CCL3, CXCL10, IL-18, PD-L1, TNFR1, ALC, bilirubin, BUN, Cr, Hct, LPS-induced IL-6 production, temp | bilirubin |
| 16 | 0.729 | CCL3, CXCL10, IL-18, PD-L1, TNFR1, ALC, BUN, Cr, Hct, LPS-induced IL-6 production, temp | CXCL10 |
| 17 | 0.729 | CCL3, IL-18, PD-L1, TNFR1, ALC, BUN, Cr, Hct, LPS-induced IL-6 production, temp | temp |
| 18 | 0.712 | CCL3, IL-18, PD-L1, TNFR1, ALC, BUN, Cr, Hct, LPS-induced IL-6 production | LPS-induced IL-6 production |
| 19 | 0.695 | CCL3, IL-18, PD-L1, TNFR1, ALC, BUN, Cr, Hct | Hct |
| 20 | 0.712 | CCL3, IL-18, PD-L1, TNFR1, ALC, BUN, Cr | TNFR1 |
| 21 | 0.729 | CCL3, IL-18, PD-L1, ALC, BUN, Cr | PD-L1 |
| 22 | 0.729 | CCL3, IL-18, ALC, BUN, Cr | ALC |
| 23 | 0.678 | CCL3, IL-18, BUN, Cr | IL-18 |
| 24 | 0.627 | CCL3, BUN, Cr | Cr |
| 25 | 0.492 | CCL3, BUN | CCL3 |
| 26 | 0.576 | BUN | N/A |

\* ALC = absolute lymphocyte count; BUN = blood urea nitrogen, plasma; Cr = creatinine, plasma; DBP = diastolic blood pressure; GCS = Glasgow coma score; Hct = hematocrit; LPS = lipopolysaccharide stimulation, ex vivo; SOFA (max) = maximum SOFA score recorded within 14 days of illness onset; temp = temperature

## Discussion

Our proteomics-driven analysis identified a distinctive molecular signature distinguishing septic and non-septic critical illnesses, including lower levels of key positive acute-phase reactants (serum amyloid A1/2, C-reactive protein, lipopolysaccharide-binding protein, and von Willebrand factor) in sepsis patients. Among all bedside variables, serum creatinine emerged as the single strongest proteome guided correlate (R² = 0.56) and remained in the final minimal model, together with BUN and CCL3. This finding reinforces accumulating evidence that sepsis associated acute kidney injury is both common and prognostically pivotal (11, 12) and suggests that real-time tracking of renal biomarkers could serve as a high-yield surrogate for broader proteomic disturbances. Taken together, these results suggest that assigning equal weights to all organ systems within the SOFA score may overlook the disproportionate diagnostic impact of renal dysfunction. Incorporating weighted or biomarker-informed adjustments can substantially improve the risk stratification in sepsis. Such refinements would align organ scoring with the underlying biology, bringing clinical assessment closer to true disease mechanisms.

Similarly, declining absolute lymphocyte counts and rising inflammatory chemokines (CCL3, IL 18) converged in our parsimonious nine variable model, echoing reports that lymphopenia and chemokine storms independently portend worse outcomes (13). By integrating these easily obtainable laboratory tests with organ-specific scores, clinicians may capture the immune-metabolic state signaled by expensive proteomic assays without needing them at the bedside.

Our results advocate a ‘proteomics-informed, clinical-first - paradigm: use discovery-phase omics to identify the physiological systems most perturbed in sepsis, and then refine conventional scores by re-weighting select components rather than adding more biomarkers. For example, creatinine and BUN could be given greater influence within a focused renal-SOFA, analogous to how lactate has been incorporated into a modified cardiovascular-SOFA (14–16). Such recalibration aligns the diagnostic effort with pathobiology, while preserving the ease of implementation.

Multi analyte panels, such as presepsin with MR proADM, achieve AUCs ≈ 0.90 in some cohorts (17), yet their incremental benefit over optimized clinical data remains uncertain. Our nine variable model attained an AUC of 0.78 using only laboratory measures, demonstrating that biomarker driven feature selection can enhance—not replace—clinical judgment. Integration within electronic health record systems would allow near instantaneous risk stratification without additional costs or turnaround time.

Key limitations include the single-center design, modest sample size, and exclusion of conventional biomarkers (CRP, procalcitonin) due to missingness. We are addressing these gaps in an ongoing validation study that will prospectively compare the focused renal-SOFA and full SOFA scores against established biomarker panels. The reliance on the Sepsis-3 definition inherently introduced circularity between predictors and outcomes, although our model ultimately employed independent variables. Finally, although pathway analyses have implicated hepatocyte acute phase signaling and PD L1–mediated immunoregulation, causal relationships require targeted mechanistic studies.

## Conclusions

A compact, BUN- and creatinine-anchored variable set, derived from unbiased omics, achieves strong diagnostic performance and lays the groundwork for next-generation mechanism aware sepsis scores that are both biologically informed and clinically pragmatic.

## Ethical Conduct of Research

This study was approved by the Penn State University Human Subjects Protection Office (IRB Protocol #15328, 7/30/2020). The studies were conducted in accordance with local legislation and institutional requirements. All participants, or their legally authorized healthcare representatives, provided written informed consent to participate in the study.

## Clinical Trials Registry

The present investigation was prospectively registered on clinicaltrials.gov (registration number NCT03146546).

## Data Availability

All data produced in the present study are available upon reasonable request to the authors

## Acknowledgments

Mass Spectrometry and Proteomics Core (RRID:SCR_017831) services and instruments used in this project were funded, in part, by the Pennsylvania State University College of Medicine via the Office of the Vice Dean of Research and Graduate Students and the Pennsylvania Department of Health using Tobacco Settlement Funds (CURE). The Pennsylvania Department of Health specifically disclaims responsibility for any analysis, interpretation or conclusion.

The Penn State Critical Illness and Research Center (RRID: SCR_026307) is a multidisciplinary consortium of investigators who share data and resources related to critical illness research within the Penn State system. The content is solely the responsibility of the authors and does not necessarily represent the official views of the University or College of Medicine.

We would like to thank Abigail Samuelsen and Ruth-Ann Brown for their assistance in curating data and collecting patient samples.

## Disclosure Statement

The authors declare no competing interests.

## Data Availability Statement

Data supporting this study are available from the corresponding author on reasonable request.

## Health and Safety

All experiments adhered to approved institutional biosafety protocols (Penn State Protocol # 202200124).

## Author Contributions

AKA and ASB conceived the study. AKA, SR, and ASB designed the study. AKA and SR developed the software. AKA performed formal analyses. AKA and ASB prepared the figures and drafted the manuscript. All authors reviewed and edited the final version. ASB supervised the project and secured funding.

## Notes

### Competing Interest Statement

The authors have declared no competing interest.

### Funding Statement

National Institute of General Medical Sciences (R35GM150695, ASB)

### Author Declarations

Human Subjects Protection Office of Penn State Hershey Medical Center gave ethical approval for this work

## References

1. Singer M, Deutschman CS, Seymour CW, Shankar-Hari M, Annane D, Bauer M, et al. The Third International Consensus Definitions for Sepsis and Septic Shock (Sepsis-3). JAMA. 2016;315(8):801–10.

2. Vincent JL, Moreno R, Takala J, Willatts S, De Mendonca A, Bruining H, et al. The SOFA (Sepsis-related Organ Failure Assessment) score to describe organ dysfunction/failure. On behalf of the Working Group on Sepsis-Related Problems of the European Society of Intensive Care Medicine. Intensive Care Med. 1996;22(7):707–10.

3. Pimienta G, Heithoff DM, Rosa-Campos A, Tran M, Esko JD, Mahan MJ, et al. Plasma Proteome Signature of Sepsis: a Functionally Connected Protein Network. Proteomics. 2019;19(5):e1800389.

4. von Elm E, Altman DG, Egger M, Pocock SJ, Gotzsche PC, Vandenbroucke JP, et al. The Strengthening the Reporting of Observational Studies in Epidemiology (STROBE) statement: guidelines for reporting observational studies. Lancet. 2007;370(9596):1453-7.

5. Subbe CP, Kruger M, Rutherford P, Gemmel L. Validation of a modified Early Warning Score in medical admissions. QJM. 2001;94(10):521–6.

6. Gardner-Thorpe J, Love N, Wrightson J, Walsh S, Keeling N. The value of Modified Early Warning Score (MEWS) in surgical in-patients: a prospective observational study. Ann R Coll Surg Engl. 2006;88(6):571–5.

7. Charlson ME, Pompei P, Ales KL, MacKenzie CR. A new method of classifying prognostic comorbidity in longitudinal studies: development and validation. J Chronic Dis. 1987;40(5):373–83.

8. Knaus WA, Draper EA, Wagner DP, Zimmerman JE. APACHE II: a severity of disease classification system. Crit Care Med. 1985;13(10):818–29.

9. Ferreira FL, Bota DP, Bross A, Melot C, Vincent JL. Serial evaluation of the SOFA score to predict outcome in critically ill patients. Jama-J Am Med Assoc. 2001;286(14):1754–8.

10. Vincent JL, de Mendonca A, Cantraine F, Moreno R, Takala J, Suter PM, et al. Use of the SOFA score to assess the incidence of organ dysfunction/failure in intensive care units: Results of a multicenter, prospective study. Critical Care Medicine. 1998;26(11):1793–800.

11. Flannery AH, Li X, Delozier NL, Toto RD, Moe OW, Yee J, et al. Sepsis-Associated Acute Kidney Disease and Long-term Kidney Outcomes. Kidney Med. 2021;3(4):507–14 e1.

12. Wang Z, Dong S, Qin Y. The Relationship Between Acute Kidney Injury in Sepsis Patients and Coagulation Dysfunction and Prognosis. Open Access Emerg Med. 2024;16:145–57.

13. Charoensappakit A, Sae-Khow K, Vutthikraivit N, Maneesow P, Sriprasart T, Pachinburavan M, et al. Immune suppressive activities of low-density neutrophils in sepsis and potential use as a novel biomarker of sepsis-induced immune suppression. Sci Rep. 2025;15(1):9458.

14. Lee HJ, Ko BS, Ryoo SM, Han E, Suh GJ, Choi SH, et al. Correction: Modified cardiovascular SOFA score in sepsis: development and internal and external validation. BMC Med. 2022;20(1):476.

15. Lee HJ, Ko BS, Ryoo SM, Han E, Suh GJ, Choi SH, et al. Modified cardiovascular SOFA score in sepsis: development and internal and external validation. BMC Med. 2022;20(1):263.

16. Lorstad S, Wang Y, Tehrani S, Shekarestan S, Astrand P, Gille-Johnson P, et al. Development of an Extended Cardiovascular SOFA Score Component Reflecting Cardiac Dysfunction with Improved Survival Prediction in Sepsis: An Exploratory Analysis in the Sepsis and Elevated Troponin (SET) Study. J Intensive Care Med. 2025;40(3):320–30.

17. Liang J, Cai Y, Shao Y. Comparison of presepsin and Mid-regional pro-adrenomedullin in the diagnosis of sepsis or septic shock: a systematic review and meta-analysis. BMC Infect Dis. 2023;23(1):288.

