## Supplementary Materials and Methods for "Diagnosing Sepsis Through Proteomic Insights: Findings from a Prospective ICU Cohort"

### **Supplementary Material**

**Supplementary Methods**

***Proteomic Analysis***

Blood samples were drawn into K2-EDTA tubes, immediately centrifuged to isolate plasma, and stored at -80°C. Prior to analysis, samples were thawed at 4°C, supplemented with HALT protease inhibitor (ThermoFisher), and processed in a single batch using MagReSyn HILIC beads (ReSyn Biosciences) in a 96-well plate format on the KingFisher Flex system. Plasma proteins underwent reduction, alkylation, and digestion on beads. Digested peptides were acidified, spiked with trypsin-digested equine myoglobin for quality control, centrifuged to remove beads, quantified by NanoDrop, and analyzed.

***Liquid Chromatography-Mass Spectrometry***

Peptides were analyzed using a Bruker TimsTOF Flex mass spectrometer coupled with NanoElute LC in dia-PASEF mode. A µ-Precolumn and PepSep C18 analytical column were used for separation with a ~20-min gradient. DIA methods included optimized isolation windows across ion mobility scans, leveraging in-silico peptide libraries (pydiAID software, (18)).

***Mass Spectrometry Data Processing***

Spectral data were processed using DIA-NN in two phases (19). Initially, an experiment-specific spectral library was generated from a subset of samples (including QC replicates). Subsequently, all samples were analyzed against this library. Search parameters included one missed cleavage, fixed C-carbamylmethylation, variable N-terminal modifications, oxidation of methionine, 15 ppm mass accuracy, and a default 1% FDR cutoff for protein identification.

***Statistical and Functional Analysis***

Differential proteomic analyses identified proteins significantly different between sepsis and critical illness non-sepsis (CINS) groups. Data underwent filtering (minimum 75% non-missing globally and per group) and K-nearest neighbor imputation. Ingenuity Pathway Analysis (QIAGEN) identified significantly dysregulated proteins (log2 fold change >1 or <–1, adjusted P<0.1) for downstream enrichment analyses.

***Proteomic-Enrichment of Clinical Predictors (Discovery Cohort)***

Using Random Forest (RF) regression models, we evaluated if differentially expressed proteins predicted clinical variables (continuous and binary). Continuous variables with R²>0 and binary variables with AUC > 0.6 were selected. Missing continuous data (<50%) were imputed using Multiple Imputation by Chained Equations (MICE). Leave-One-Out Cross-Validation (LOOCV) with 10 repetitions assessed model stability.

***Validation Cohort***

A refined RF model was trained using the Discovery cohort's selected variables, applying Youden's J statistic for threshold optimization. The model was validated on an independent cohort (Validation cohort, n=59) using ROC analyses, repeated 10 times for result stability.

***Recursive Feature Elimination***

Recursive Feature Elimination (RFE) with RF iteratively identified the minimal optimal set of clinical variables predictive of sepsis, removing the least important variable at each step until predictive accuracy was optimized.

**Supplementary Figures**

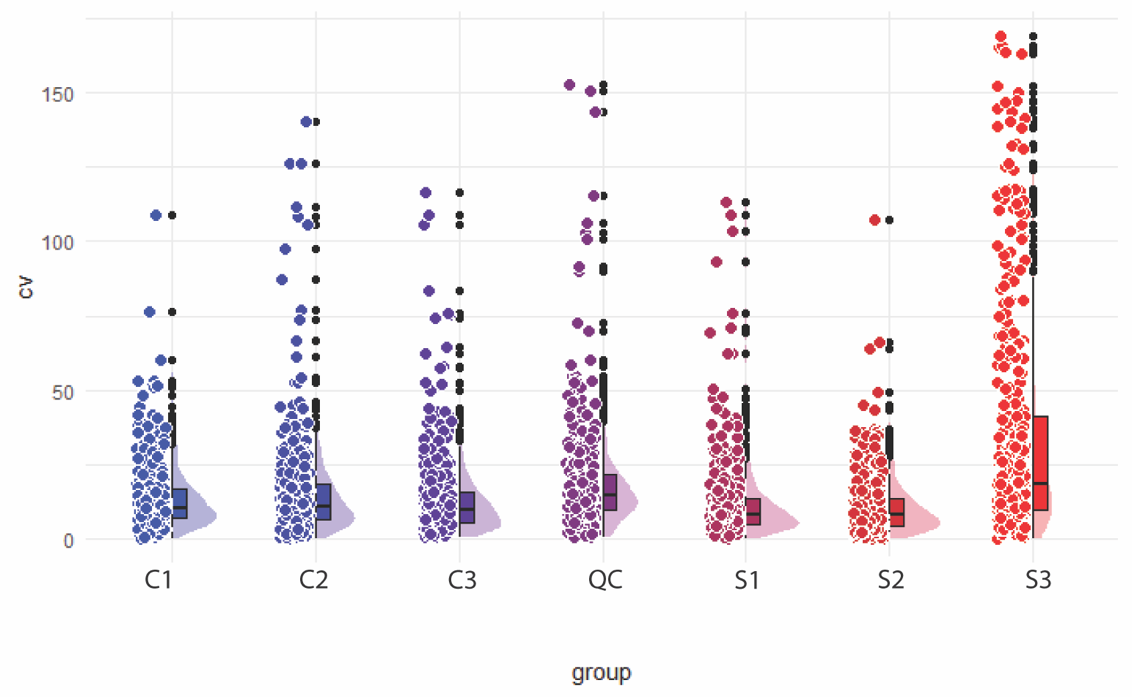

**Fig S1. Raincloud plot showing coefficient of variation** **between triplicate measurements from the same patients.** Displayed are three critically ill and non-septic patients (left, C1-C3), three critically ill and septic patients (right, S1-S3) and quality control sample (center, QC).

**Supplementary Tables**

**Supplementary Table S1: Clinical Features used in Discovery Analysis**

|  | **Variable** | **Variable type** | |
| --- | --- | --- | --- |
| **Demographics/**  **Anthropometrics** | Age | Numeric | Continuous |
|  | Sex | Numeric | Binary |
|  | Race | Numeric | Categorical |
|  | Body Mass Index | Numeric | Continuous |
|  | Skeletal muscle index | Numeric | Continuous |
| **Clinical Presentation and Severity Scores** | Presence of sepsis | Numeric | Binary |
|  | Presence of shock | Numeric | Binary |
|  | APACHE II score | Numeric | Discrete |
|  | Charlson Comorbidity Index | Numeric | Discrete |
|  | Glasgow Coma Score | Numeric | Discrete |
|  | Sequential Organ Failure Assessment (SOFA) Score, day 1 of illness | Numeric | Discrete |
|  | Primary organism type | Numeric | Categorical (fungal, bacterial, viral) |
|  | Maximum SOFA score, days 1-15 | Numeric | Discrete |
| **Hematologic and Biochemical Laboratory Values** | Absolute leukocyte count | Numeric | Continuous |
|  | Absolute lymphocyte count | Numeric | Continuous |
|  | Absolute monocyte count | Numeric | Continuous |
|  | Hematocrit | Numeric | Continuous |
|  | Platelet count | Numeric | Continuous |
|  | Bicarbonate concentration, serum | Numeric | Continuous |
|  | Aspartate aminotransferase, serum | Numeric | Continuous |
|  | Blood urea nitrogen, serum | Numeric | Continuous |
|  | Albumin, serum | Numeric | Continuous |
|  | Bilirubin, serum | Numeric | Continuous |
|  | Creatinine, serum | Numeric | Continuous |
|  | C-reactive protein, serum | Numeric | Continuous |
|  | Lactate, serum | Numeric | Continuous |
|  | Procalcitonin, serum | Numeric | Continuous |
| **Immunologic and Proteomic Markers** | CCL3, plasma | Numeric | Continuous |
|  | CCL7, plasma | Numeric | Continuous |
|  | Interleukin-10, plasma | Numeric | Continuous |
|  | PD-L1, plasma | Numeric | Continuous |
|  | CXCL9, plasma | Numeric | Continuous |
|  | CXCL10, plasma | Numeric | Continuous |
|  | Fas Ligand, plasma | Numeric | Continuous |
|  | TRAIL, plasma | Numeric | Continuous |
|  | Interleukin-8, plasma | Numeric | Continuous |
|  | Interleukin-18, plasma | Numeric | Continuous |
|  | Leptin, plasma | Numeric | Continuous |
|  | TNF receptor 1, plasma | Numeric | Continuous |
|  | Interferon-gamma, plasma | Numeric | Continuous |
|  | Interleukin-6, plasma | Numeric | Continuous |
|  | Tumor necrosis factor, plasma | Numeric | Continuous |
|  | Ex vivo, Lipopolysaccharide-stimulated Interleukin-6 concentration | Numeric | Continuous |
|  | Ex vivo, Lipopolysaccharide-stimulated TNF concentration | Numeric | Continuous |
|  | Ex vivo, Anti-CD3/anti-CD28-stimulated interferon-gamma concentration | Numeric | Continuous |
|  | Ex vivo, PMA-stimulated interferon-gamma concentration | Numeric | Continuous |
| **Physiologic Parameters** | SpO2 | Numeric | Continuous |
|  | Temperature | Numeric | Continuous |
|  | PaO2:FiO2 ratio | Numeric | Continuous |
|  | Minimum systolic blood pressure in 24 hours | Numeric | Continuous |
|  | Minimum diastolic blood pressure in 24 hours | Numeric | Continuous |
|  | Minimum mean arterial pressure in 24 hours | Numeric | Continuous |
|  | Highest respiratory rate in 24 hours | Numeric | Discrete |
|  | Mechanical ventilation | Numeric | Binary |

**Supplementary Table S2.** Strength of the association between the top 12 identified proteins and the clinical variables in the prediction of sepsis as an outcome (n=10 iterations).

| **Clinical Feature** | **R^2^** |
| --- | --- |
| Cr | 0.558 |
| PD-L1 | 0.387 |
| IL-8 | 0.363 |
| albumin | 0.238 |
| TNFR1 | 0.205 |
| Hct | 0.199 |
| BUN | 0.169 |
| GCS | 0.155 |
| SOFA | 0.154 |
| APACHE II score | 0.152 |
| CCL3 | 0.149 |
| IL-6 | 0.141 |
| SOFA (max) | 0.141 |
| lactate | 0.117 |
| ALC | 0.113 |
| CXCL10 | 0.104 |
| IL-10 | 0.088 |
| LPS-induced IL-6 production | 0.058 |
| age | 0.052 |
| IL-18 | 0.032 |
| LPS-induced TNF production | 0.021 |
| bilirubin | 0.019 |
| DBP | 0.015 |
| temperature | 0.005 |

### * ALC = absolute lymphocyte count; BUN = blood urea nitrogen, plasma; Cr = creatinine, plasma; DBP = diastolic blood pressure; GCS = Glasgow coma score; Hct = hematocrit; LPS = lipopolysaccharide stimulation, ex vivo; SOFA (max) = maximum SOFA score recorded within 14 days of illness onset.
